# Autocrine and paracrine interferon signaling as ‘ring vaccination’ and ‘contact tracing’ strategies to suppress virus infection in a host

**DOI:** 10.1101/2020.12.09.20246777

**Authors:** G. Michael Lavigne, Hayley Russell, Barbara Sherry, Ruian Ke

## Abstract

The innate immune response, particularly the interferon response, represents a first line of defense against viral infections. The interferon molecules produced from infected cells act through autocrine and paracrine signaling to turn host cells into an antiviral state. Although the molecular mechanisms of IFN signaling have been well characterized, how the interferon response collectively contribute to the regulation of host cells to stop or suppress viral infection during early infection remain unclear. Here, we use mathematical models to delineate the roles of the autocrine and the paracrine signaling, and show that their impacts on viral spread are dependent on how infection proceeds. In particular, we found that when infection is well-mixed, the paracrine signaling is not as effective; in contrast, when infection spreads in a spatial manner, a likely scenario during initial infection in tissue, the paracrine signaling can impede the spread of infection by decreasing the number of susceptible cells close to the site of infection. Furthermore, we argue that the interferon response can be seen as a parallel to population-level epidemic prevention strategies such as contact tracing or ring vaccination. Thus, our results here may have implications for the outbreak control at the population scale more broadly.

## 1 Introduction

The innate immune response provides critical protection against pathogen invasion of humans and other animals prior to establishment of adaptive immunity. It relies on multiple cytokines, chief among them being interferons (IFNs), a large, diverse family of signaling proteins that together induce a protective response [1]. The importance of IFN in the defense against viral infections is demonstrated by the fact that essentially all viral pathogens have developed mechanisms to interfere with or suppress the host IFN response [2, 3, 4]. Indeed, viral evasion of the IFN response strongly determines the rate of viral replication, the success of transmission and infection establishment in new hosts [5] and the range of species infected [6]. The capacity to inhibit the IFN response determines species tropism for human immunodeficiency virus [7], dengue virus [8], rotavirus [9], measles virus [10], and influenza virus [11]. Interestingly, multiple lines of recent evidence show that severe symptoms and life threatening disease from SARS-CoV-2 infection is linked to inhibition or dysfunction of antiviral response mediated by IFN signaling or inborn deficiency in IFN immunity [12, 13, 14, 15].

The IFN response is commonly described by its two components: first, viral induction of IFN, and second, IFN induction of antiviral genes [16]. Upon infection, viral RNAs or DNAs are detected by the cell triggering a signaling cascade that results in the production of Type I IFNs [17, 18]. These IFN molecules are then secreted and bind to surface receptors located on the cell membrane. IFN binding to the surface of the cell from which it is produced is referred to as autocrine signaling, whereas binding to the surface of any other cell is referred to as paracrine signaling. This binding initiates a series of signaling events that ultimately result in the production of Interferon Stimulated Genes (ISGs), the expression of which repress viral replication in the cell at multiple steps [19]. In an uninfected cell, binding of IFN to its receptor and subsequent IFN signaling renders the cell refractory to viral infection, while in an infected cell, this signaling can suppress viral replication and decrease release of viral progeny from the cell. An elegant analysis of the virus-induced IFN response at the single cell level demonstrated that paracrine signaling early in infection shapes the overall IFN response [20]. However, the inflammatory response elicited by IFN can have deleterious effects on the host if uncontrolled [21, 22, 12, 15].

Although the molecular mechanisms of IFN signaling have been well characterized, the systems-level properties arising from the individual host cell response, particularly how the host cells collectively stop viral infection at the site of exposure before the infection becomes systematic remain unclear. To address these questions, we use modeling approaches to understand how IFN signaling can stop early infection (e.g. at the site of initial entry) before adaptive immunity is developed. Previous modeling of virus infection and the IFN response has focused on the role of IFN response after the infection becomes systematic and used ordinary differential equations (ODEs) [23, 24, 25, 26]. ODE models necessarily include the implicit assumption that the host is treated as a single well-mixed compartment, and thus they neglect the spatial structure of infection. Influenza infection, for example, starts at the epithelial lining of the upper respiratory tract, which is an inherently a spatial process [27]. Therefore, to investigate the interaction between virus and the IFN response early in infection, a spatially explicit model is most appropriate. We previously modelled the spread of virus infection and the effectiveness and robustness of IFN signaling among host cells using a network approach [28, 29]. however, the explicit roles of autocrine and paracrine signaling in suppressing virus spread are not clear.

Here, we develop various models with or without explicitly considering the impact of the spatial arrangement of cells to examine the roles of autocrine and paracrine IFN signaling. We first show that, in well-mixed ODE models, autocrine signaling can impact the course of infection by inhibiting virus production from already infected cells, whereas paracrine signaling has negligible impact on the growth of viral load during early infection when target cells are abundant. In contrast, in models explicitly considering spatial spread, IFN paracrine signaling can stop viral infection by segregating susceptible cells from areas of infection with an insulating layer of protected cells. This strategy parallels the control strategies of “ring vaccination” and “contact tracing” in epidemiology and outbreak control which aim to stop spread of infection by targeting the most at-risk individuals [30, 31, 32].

## 2 Methods

### 2.1 A non-spatial model of well-mixed viral infection

We first develop a model of viral infection with IFN signaling using ordinary differential equations (ODEs). In this approach, we assume that cells, viruses and IFN are well mixed and thus spatial structure is not considered. Such models have been well established by previous work on in vivo models of virus-immune interaction during systemic infection [23, 25]. The equations of our model are as follows:

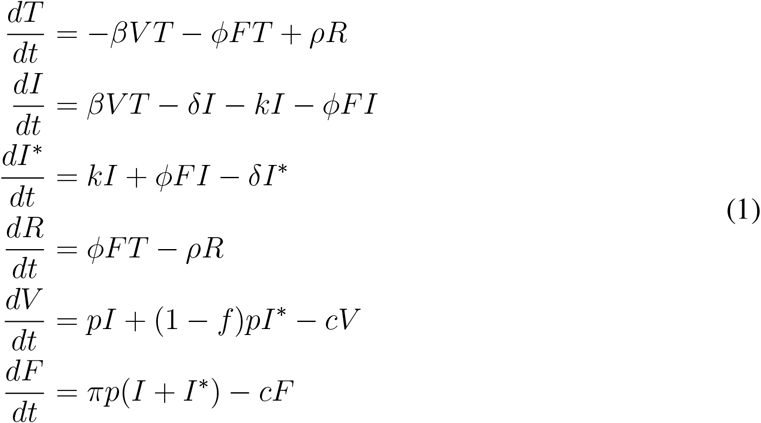

In this model (see Figure 1(a) for a schematic), cells are categorized into one of four states: uninfected target cells *T*, productively infected cells *I*, infected cells that are in an antiviral state *I*^∗^ and refractory cells *R*. Uninfected cells are infected by virions *V* at rate *β* or become refractory to infection through paracrine signaling of IFN (*F*) at rate *ϕ*. Binding of IFN molecules to IFN receptors on infected cells (*I*), including both autocrine and paracrine IFN signaling, may trigger an antiviral response in those cells, such that virus production is inhibited or reduced [24]. We model the impacts of autocrine and paracrine signaling using two separate terms, i.e. *kI* and *ϕFI*. Note that *F* in our model represents the ambient concentration of unbound IFN (under the assumption of homogeneous concentration of IFN). We assume that autocrine signaling occurs independent of the ambient IFN concentration, because once produced from infected cells, IFNs preferentially bind to the producing cell due to proximity. The transition towards an antiviral state due to autocrine signaling is thus modeled by *kI*, i.e. independent of ambient IFN concentration. In contrast, the rate of transition due to the paracrine signaling is modeled to be dependent on the IFN concentration with the term *ϕFI*.

**Figure 1:**
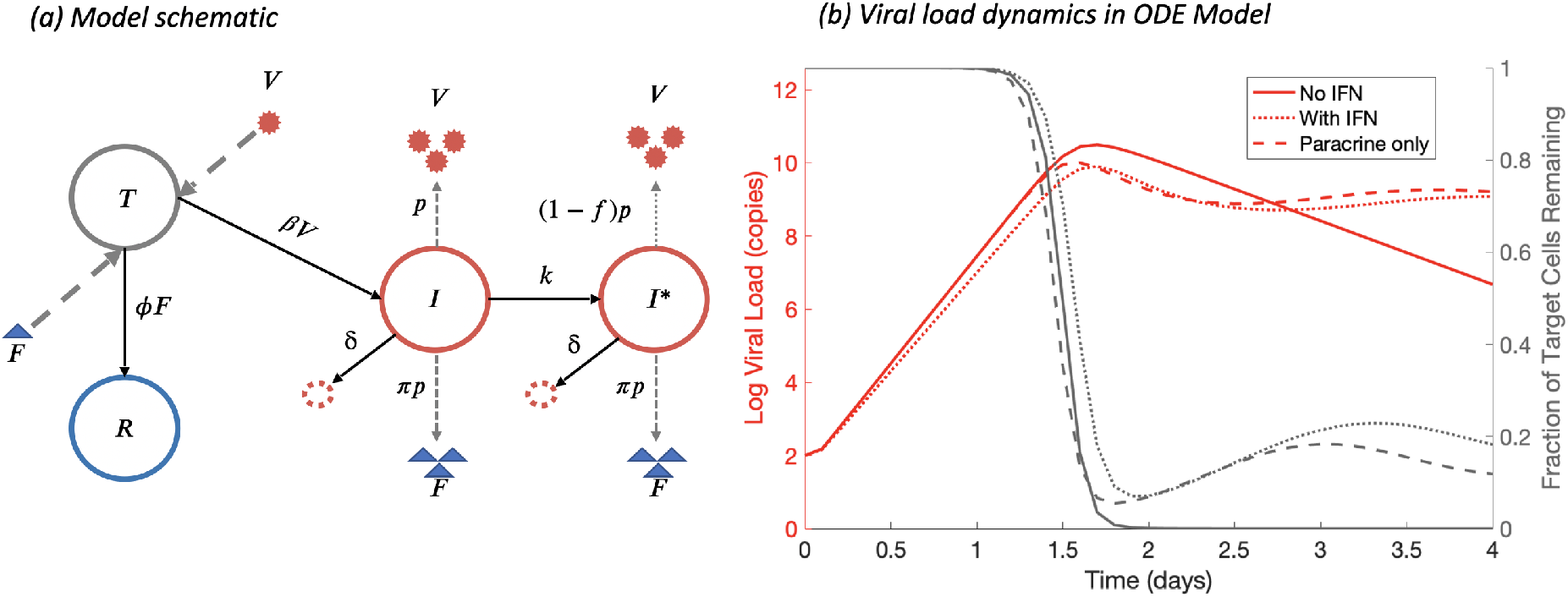
Schematic diagram of the viral infection dynamics and the IFN response and simulations of the corresponding ODE model. **(a)** Schematic diagram with parameters in the model. Solid arrows indicate transition of cells from one state to another; dashed arrows indicate the production or binding of viruses and IFNs from cells. **(b)** Simulations of ODE model without IFN, with IFN, and with only paracrine signaling demonstrate the effects of different parameter regimes on the growth of viral load. Simulation of a model with the paracrine signaling alone (*k* = 0, dashed lines) shows no notable effect on the initial exponential growth rate compared to the no-IFN case (*k* = 0 and *π* = 0, solid lines), while both autocrine and paracrine signaling together can slow the growth of viral load (dotted lines).

We assume that infected cells (both *I* and *I*^∗^) die at the same per capita rate *δ*. Refractory cells remain protected for an average time of 1/*ρ* before returning to the susceptible state, i.e. becoming target cells again. Infected cells, *I*, release viruses at rate p, whereas infected cells at an antiviral state, *I*^∗^, release virions at a reduced rate (1 −*f*)*p*, where *f* is the fraction of reduction. For simplicity, we further assume that both *I* and *I*^∗^ cells release IFNs at rate *πp* and that viruses and IFNs are cleared at per capita rate *c*. Note that since the time scale of the dynamics of IFNs is much faster than the time scale of dynamics of the cells, we can make the quasi-equilibrium assumption for the concentration of IFN and then the level of IFNs are related to infected cells as 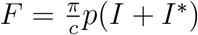. Therefore, if IFN is cleared in the system at a rate different from *c*, the level of IFN can be compensated in the system by changing the value of *π*.

### 2.2 A Spatial Model Viral Infection with IFN Signaling

We next develop a partial differential equation (PDE) model of viral infection and IFN response. This model explicitly considers the spatial arrangement of cells, virions, and IFNs, thus more accurately representing the dynamics of infection in an epithelial tissue. We assume that susceptible cells *T* are arranged on a 1-dimensional space with spatial variable *x ∈* [0, *L*] with a uniform initial density *T*_0_. Viruses and IFNs can diffuse to nearby locations, in contrast to the ODE model where viruses and IFNs are assumed to instantaneously be evenly distributed once produced. Virions and IFNs diffuse across the spatial domain with diffusion coefficients *D*_*V*_ and *D*_*F*_, where we take *D*_*F*_ >> *D*_*V*_ since IFNs are much smaller than virions and therefore diffuse at a much greater rate [33, 34]. These diffusion parameters determine the characteristic length scales on which IFNs and virions will be active [35]. The initial conditions are taken to be such that the domain is populated only with target cells at a constant density and a single infected cell at the position *x* = 0, which is achieved using a Dirac delta distribution *δ*_0_(*x*). The boundary conditions are taken to be homogeneous Neumann at *x* = 0 to represent reflective symmetry of the spread of infection, and homogeneous Dirichlet at far-field *x* = *L*. The equations of the model are as follows:

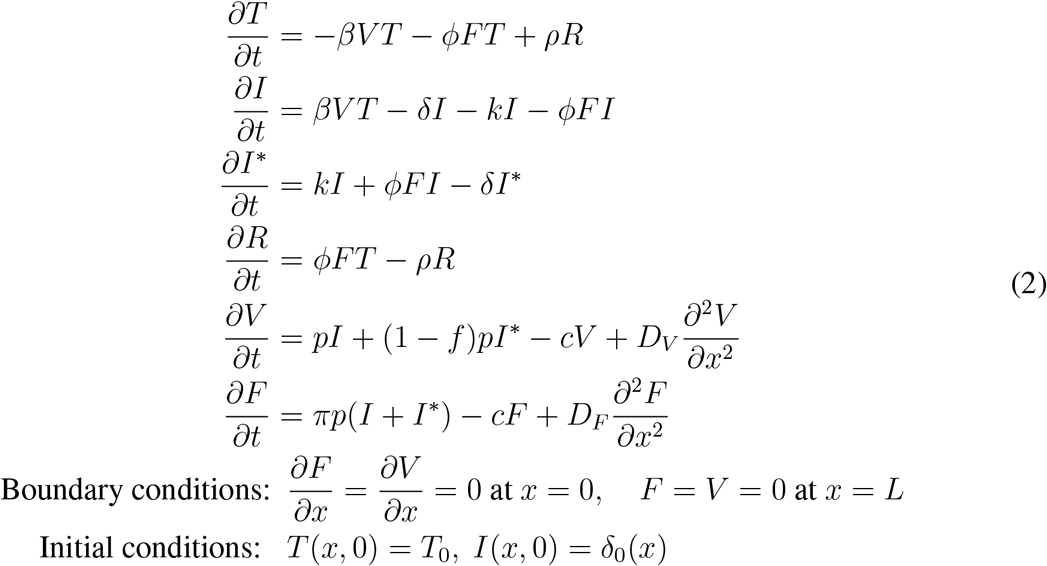

### 2.3 A Cellular Automata Model

We lastly develop a 2D Cellular Automata model (CA) to model the spatial progression of viral infection with IFN signaling. The CA framework allows us to consider the spatial infection spread governed by a stochastic process. By developing a CA model, we can more accurately depict the nature of early viral infection in epithelial tissue

In our CA model, each individual epithelial cell is tracked explicitly as a grid point on a stationary *N* ×*N* lattice. Cells interact locally with other cells near to themselves based on predefined rules for the production and diffusion of virions and IFN particles. A cell can be in any of five states: healthy, exposed, productively infected, protected, or dead. The CA is initialized with a single infected cell located at the center of the grid of otherwise healthy target cells. Furthermore, virion and interferon particles are not explicitly modeled agents, but rather we consider their production, diffusion, and binding to recipient cells to occur within the duration of a single iteration of the CA. This choice allows us to take large time steps and is less costly than explicitly modeling the random walk of each particle. More detailed specifications of the CA model can be found in Supplementary Materials.

## 3 Results

### 3.1 The roles of autocrine and paracrine IFN signaling in a non-spatial well-mixed infection

We first constructed a model (see Fig. 1a for a schematic) and analyzed the roles of autocrine and paracrine IFN signaling using ordinary differential equations (ODEs) (see Methods). To understand the impacts of autocrine and paracrine signaling on the virus dynamics after initial viral exposure, we calculated the basic reproductive number *R*_0_ of the virus using the Next Generation Matrix technique [36]. Note that *R*_0_ = 1 is the threshold for establishment of infection, and viral population only grows when *R*_0_ > 1. Thus, for an effective innate immune response to halt viral infection, *R*_0_ has to be less than 1. For the above model we find:

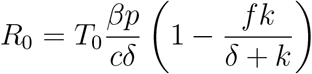

This expression shows that the reduction of *R*_0_ due to autocrine signaling is 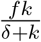, where f is the inhibition of virus production due to the cellular antiviral response and *k*/(*δ* + *k*) is the probability that an infected cell becomes antiviral by the autocrine pathway before cell death occurs *In vitro* experiments suggest that the fraction of infected cells that successfully enter an antiviral state is in general low [37, 38, 39], i.e. *k*/(*δ* +*k*) is much less than 1. If this observation is consistent with IFN response *in vivo*, then our results suggest that autocrine signaling has limited impact on stopping viral infection during initial stage of infection.

Importantly, we found that the parameters governing paracrine IFN signaling (i.e. *ϕ,π*) do not appear in the expression for *R*_0_, i.e. paracrine signaling alone does not change the infection threshold. Therefore, the ODE model makes the surprising prediction that when cells, viruses and IFN are well mixed (as assumed in our ODE model and other models [25, 23], paracrine signaling has a negligible role in halting infection during early infection when the number of target cells are abundant. We further performed simulations of the model (see Table S1 for the parameter values used for simulation) to compare the viral dynamics with and without paracrine IFN signaling (Fig. 1(b)). In agreement with the analytical derivation for *R*_0_, we found that IFN paracrine signaling has negligible impact on the viral load during initial exponential growth period. This is true even for very large (biologically unrealistic) values of *π* (Figure S1). We found that IFN-mediated protection of target cells is only able to affect the course of infection after some period of viral growth once infected cell concentration, and thus IFN concentration, rises to a sufficiently high level that there is a notable impact on protecting target cells and infected cells. The peak viral load is decreased by approximately 1/(1 + *π*)-fold and the time to peak viremia is relatively insensitive to changes in *π* (see analytical approximations in Supplementary Materials). This nominal decrease in the time to peak viremia is a consequence of the accelerated target cell depletion due to IFN signaling to uninfected cells.

Overall, our results show that when cells, viruses and IFNs are well-mixed (no spatial segregation is considered), autocrine signaling may have limited impact on the infection dynamics when a small fraction of cells turn on an antiviral response, and paracrine signaling has no impact on the infection dynamics during early infection.

### 3.2 A spatio-temporal PDE model of viral infection with IFN response

For almost all respiratory and enteric viral infections, the site of initial infection and viral replication is epithelial tissue, which is characterized by a monolayer structure [27]. Due to local diffusion of viral progeny over the epithelium, a virion is highly likely to infect one of a small number of neighboring cells rather than having an equal probability of infecting any target cell, as is the implicit assumption in an ODE model of viral infection.

To incorporate the spatial structure of host cells, we constructed a PDE model (see Eqns. 2 and Methods). We then simulated the PDE model with and without IFN signaling (Fig 2; see Table S2 for the parameter values used for simulation). In the absence of the signaling (*π* = *k* = *f* = 0; Fig 2a), the solution of the PDE model exhibits a traveling wave solution (called the infection wave below). Analyzing the PDE model, we found that a front of infected cells propagates through healthy epithelium with a constant velocity, *v*^∗^ (see Methods). An approximate expression for *v*^∗^ is as follows (see supplementary materials):

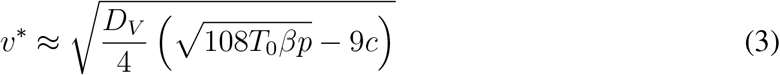

**Figure 2:**
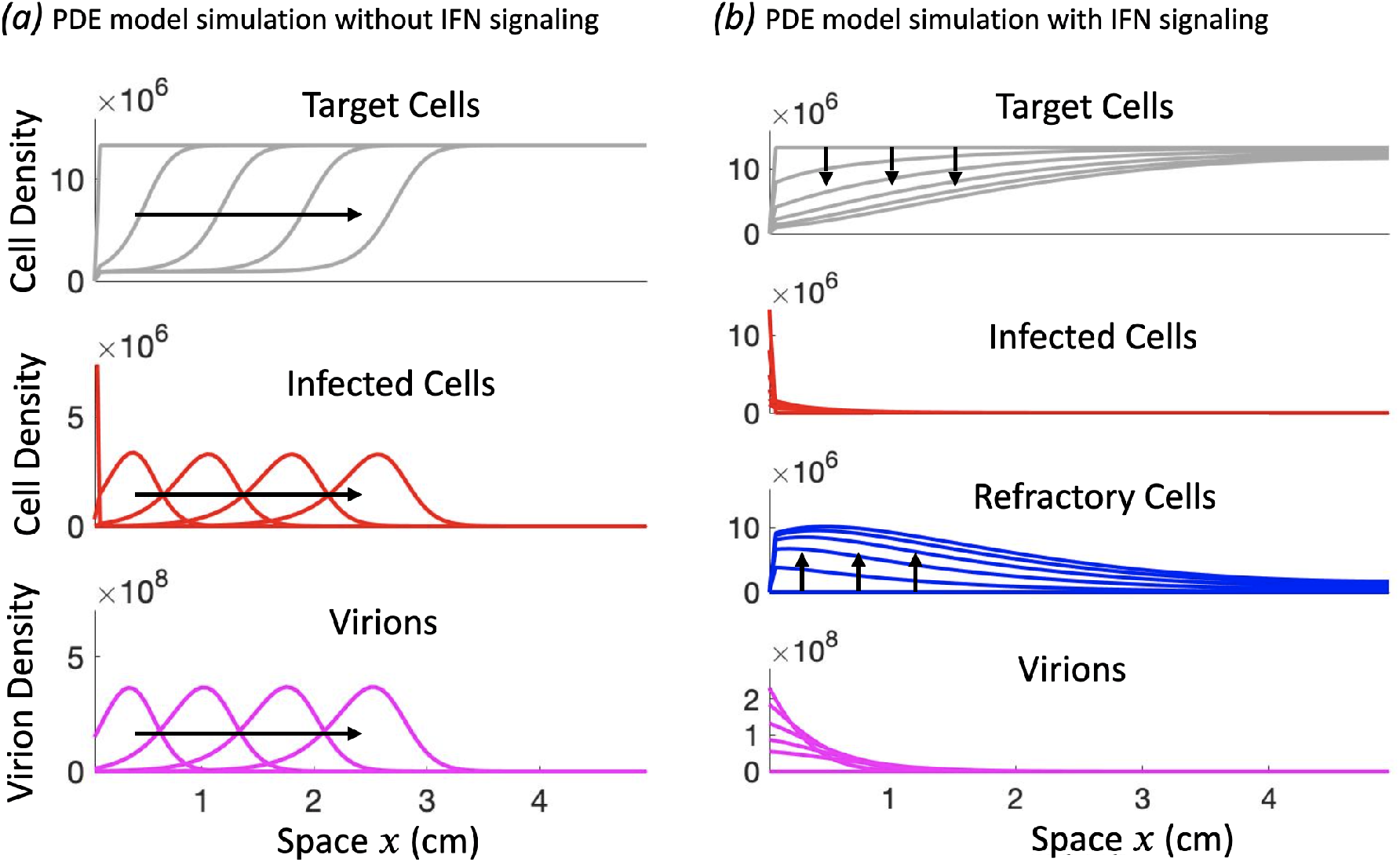
Model simulations show that paracrine IFN signaling strongly interferes with the spatial spread of infection. Shown is the solution of the model system at successive time points, with arrows indicating the direction of progression with time. **(a)** A representative simulation of the PDE model with *k* = *π* = 0 (no IFN), exhibiting traveling wave behavior initiated from a single nexus of infected cells at position *x* = 0. The infection travels an equal distance between successive times, demonstrating constant speed of spread. **(b)** A representative simulation of the PDE model with cell protection included (*π* ≠ 0) showing how IFN signaling can stop the spread of infection by depleting target cells. The distribution of virions and infected cells can be seen to remain localized to the left side of the domain, as the rapid depletion of susceptible cells (as a result of increases in refractory cell population) in the domain prevent the infection from establishing a traveling wave.

This expression shows that the spread of infection is driven primarily by the production (*p*) and diffusion (*D*_*V*_) of virions, the infection of target cells (*β*) and the density of available target cells leading the front of the infection wave, i.e. *T*_0_. The IFN signaling has the effect of both decreasing the production of virions (*p*) and decreasing the number of cells susceptible to infection (*T*_0_), and thus it can in principal slow the spread of infection.

We then simulated the model with or without the autocrine and/or the paracrine signaling. With the inclusion of IFN paracrine signaling, we find that target cells at the front of infection are more likely to become refractory than infected due to the high diffusivity of IFN relative to virions (Fig. 2b and 3a). This causes target cell density leading the front of infected cells to decrease as the number of refractory cells rises. As the infection continues to spread, the IFN level becomes high enough leading to the depletion of target cells (*T*_0_ in Eqn. ((3))), which in turn impedes the spread of infection. Therefore, the PDE model predicts that paracrine signaling can have a strong impact on the spatial spread of virus infection by protecting cells at the front of infection.

In the absence of paracrine IFN signaling (Fig. 3b), we found that surprisingly, the observed traveling wave speed of the infection does not depend strongly on the strength of autocrine signaling, i.e. the value of the autocrine parameter *k*. This is because the speed of spread is mostly driven by virus production from cells at the wave front. These infected cells are unlikely to be in an antiviral state, because of the waiting time (on average 1/*k* days) for that to occur. Thus, the results from the PDE model is in a sharp contrast to the results form the ODE model, with respect to the roles of the autocrine and the paracrine signaling on preventing the growth of infection than paracrine signaling.

**Figure 3:**
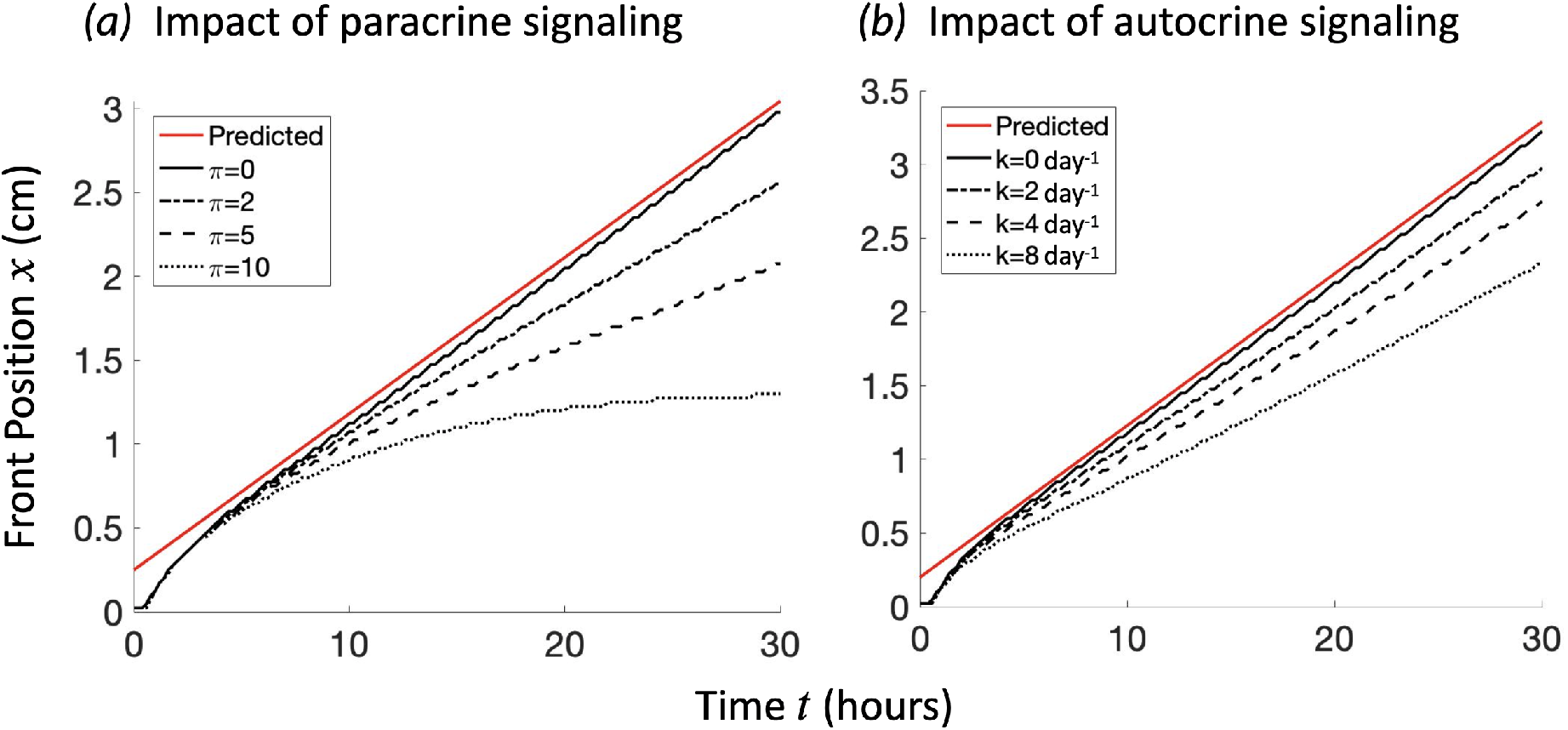
Comparison of the impacts of paracrine signaling and autocrine signaling on viral spatial spread. Shown is the position *x*(*t*) of the infection front over time for various values of the IFN production parameter *π* while keeping *k* = 0 and the autocrine-mediated transition rate *k* while keeping *π* = 0. Here we define the front position *x*(*t*) to be the position that 1% of total cells are infected, i.e. *I*(*x*(*t*), *t*) = 0.01*T*_0_. **(a)** Sufficiently strong paracrine signaling, e.g. setting *π* = 10 and *k* = 0/day, leads to halting the spread of infectious front. **(b)** Strong autocrine signaling, e.g. setting *π* = 0 and *k* = 8/day, impact modestly on the speed of the viral spread. The red lines show the predicted front position given the analytically derived wave speed in Eqn. (3).

### 3.3 A Stochastic Cellular Automata Model of the IFN response to Viral infection

The analysis of the PDE model above identified important roles of IFN paracrine signaling on the spatial and temporal dynamics of virus infection. However, the deterministic nature of the PDE model neglects the inherent stochasticity present in the early stages of viral infection by considering a continuous real-valued density of virus and IFNs rather than individual particles. Furthermore, being a parabolic system of PDEs, the densities of virus and IFN, once produced, become instantaneously non-zero everywhere in the domain. Thus, the PDE model predicts that virus infection continues in locations far away from the initial site of infection over long period of time irrespective of how strong the IFN response is (Figure S2).

Here, to understand the spread of infection in the presence of IFN signaling in a more realistic setting, we constructed a 2D cellular automaton (CA) model, similar to previous works [40, 33]. See the Methods and supplementary materials for detailed description.

We first simulated the CA model assuming there is no IFN produced (Fig. 4a). In this case, the area of infection spreads radially. In the absence of IFN, the number of infected cells increases roughly quadratically with time, suggesting that the speed of infection spread is constant (# cells infected can be approximated by the area of infection *πr*^2^ = *π*(*v*^∗^*t*)^2^), which agrees with our analysis of the PDE model. We find the growth to be sub-quadratic in the presence of effective IFN signaling, which we observe as the decrease in the slope in the log-log plots of Fig. 4 as IFN production increases. When IFN particles are produced at a low level, the infection spreads roughly the same distance as the `ifn_p`rod = 0 case (`ifn_prod` = 1 in Fig. 4b), though a lot of cells are protected by the IFN paracrine signaling (compare log-log plots in Fig. 4b with Fig. 4a). When IFN production increases further (`ifn_prod` = 5), target cells at the boundary of the front are more likely to be protected by IFN binding before becoming infected, leading to irregular spread of infection that depends heavily on virions diffusing a large distance before contacting a susceptible cell. Thus, virus spread becomes more stochastic as the spread of infection depends on rarer and rarer events (Fig. 4b). When IFN production is sufficiently large (`ifn_prod` = 20; Fig. 4d), all target cells near to infected cells are rapidly protected, leaving the virions produced each time step very unlikely to find a susceptible cell. In this way, an insulating layer of protected cells makes the continued spread of infection highly unlikely. The stronger the amount of IFN production, the thicker this layer of protected cells becomes, decreasing the probability of a virion reaching the healthy cells on the other side.

**Figure 4:**
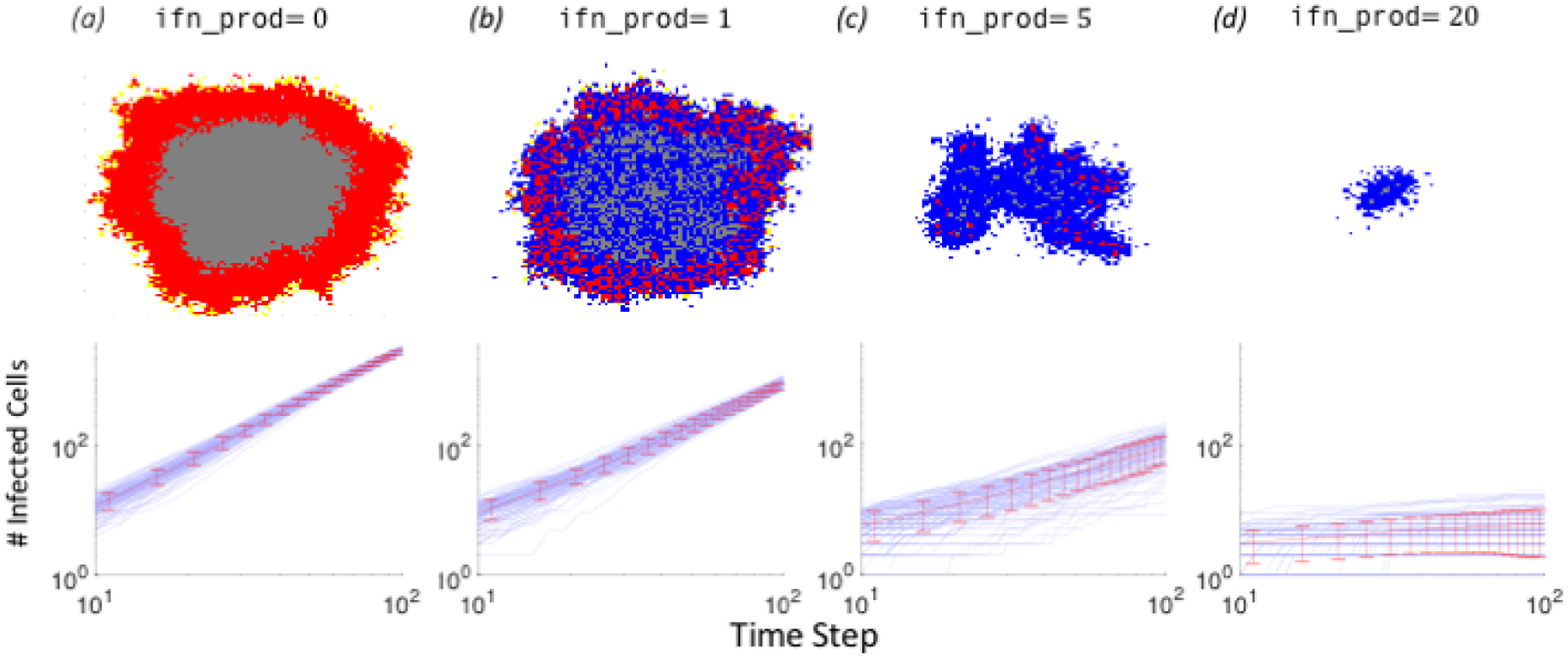
High production of IFN (ifn_prod) interferes with or stops the spread of infection by insulating infected cells from target cells with a layer of refractory cells. Top row shows example of simulations of the cellular automata (CA) model at 100 time steps on a 100 ×100 grid. Bottom row shows log-log plots of the number infected cells verses time for 100 independent simulations of CA with mean and standard deviation plotted in red. The approximate linear trend shown on the log-log plots suggests that the number of infected cells fits roughly a power law in time, *I*(*t*) ∝ *t*^*γ*^, where *γ* is the slope of the trend line. For `ifn_prod` = 0 and 1, *γ* is approximately 2, consistent with a traveling wave solution of the PDE model. *γ* decreases as `ifn_prod` increases. In the images of the CA grid, red, blue and gray dots indicate infected cells, refractory/protected cells and dead cells, respectively; Target cells are left white.

The results of the CA model corroborate our fundamental observations from the PDE model —that infection spreads at a constant rate when no IFN is present and that IFN signaling can slow and stop the spread of infection by decreasing the availability of susceptible cells in areas that are close to the site of infection. This pattern is reminiscent of the “ring vaccination” strategy in epidemiological control [31, 32, 30] where an infectious disease outbreak can be effectively controlled by vaccinating those individuals who are close to or highly likely to contact infected individuals. Furthermore, the CA model exhibits stochastic behaviors that are inadmissible in deterministic models, highlighting infection establishment during initial infection may be a stochastic event. Overall, we find that by modeling the infection and immune signaling process as spatially dependent reveals how IFN signaling can halt the spread of infection on short time and length scales by isolating infectious units from susceptible target cells.

## 4 Discussion

Using a variety of models, we demonstrated here the mechanisms by which the autocrine and the paracrine IFN signaling stop an infection before it becomes systematic. Particularly, we showed that when there exists spatial structure in host target cells (a likely scenario especially in epithelium during initial infection), the IFN response can halt an infection by rapidly inducing an anti-viral state in susceptible cells close to infected cells, thus inhibiting the ability of the infection to spread. This is likely one important mechanism by which IFN signaling is effective in suppressing early infections in epithelial tissues. These results may have important implications towards understanding the impact of early IFN response on viral dynamics and its long term role in defining disease outcomes of acute infections, such as SARS-CoV-2 infection [12, 15]. Furthermore, as we argue below, the way the IFN response controls virus infection in a host is reminiscent of the “ring vaccination” and “contact tracing” strategies in epidemiological control. Quantitative understandings of the innate immune response may provide new insights for developing effective control strategies at the epidemiological scale.

Our work clarifies the roles of autocrine and paracrine signaling and quantifies their impacts on the initiation and spread of viral infection in different host cell environments. When there exists a strong host-cell spatial structure where virion and IFN activities are restricted to locations near to where they are produced as considered in the PDE and CA models, the impact of paracrine signaling in shaping the progression of spreading infection becomes remarkably strong due to its ability to act locally. This is a likely scenario for initial stage of infection where the number of infected cells are low and infection often occurs in a restricted area of tissue epithelial cells. However, when infection occurs in an environment where spatial structure of host cells is not important, e.g. during later stage of respiratory infections where the virus population is already very large and immune response become systematic or during infections in the blood where host cells move around and contact each other, our ODE model predicts the role of autocrine signaling to be much more important than that of paracrine signaling in stymieing viral growth during early infection. Therefore, our work suggests that the two mechanisms of IFN signaling are likely complimentary to one another, though one may be more or less impactful than the other depending on the context of the infection.

Previous modeling works using ODEs predicted that the innate immune response has a strong impact on viral dynamics close to or after peak viremia [23, 24, 25]. Potent IFN response decreases the peak viral load, and decreases in IFN levels and consequently increases in target cell numbers are important to explain viral load dynamics after viral peak. Here, by explicitly considering the spatial structure of viral spread and the innate immune response, we show that potent innate immune response is able to strongly act on viral spread to slow down or even stop the virus spread during early infection. This again, highlights the important role of the IFN response throughout the infection before adaptive immune response is developed. Recently, it was hypothesized that early stochastic events in viral mutation and innate immune response during influenza infection may have long term impact on infection outcomes and disease severity [41]. In SARS-CoV-2 infection, the development of severe disease symptoms is likely due to suppression of the early antiviral response mediated by IFNs and consequently excess production of proinflammatory cytokines [12, 15]. We believe that the model framework we proposed here, together with the well-mixed approaches such in Refs. [23, 24, 25, 26], will be crucial to test the role of and quantify the impact of the IFN response during early acute infections, such as influenza and SARS-CoV-2, and how that may impact on disease outcome.

Our conclusions about how IFN response stops spatial viral spread are consistent with many lines of experimental observations. For example, it is shown in Ref. [42] that paracrine IFN signaling was able to arrest the spread of infection in a monolayer by a rapid induction of down-stream immune factors in proximal cells. Another example comes from chronic HCV infection of the liver, where the infection is highly spatially inhomogeneous, exhibiting clusters of in-fected hepatocytes surrounded by uninfected cells in which expressions of the IFN stimulated genes are high [43]. This emphasizes that IFN signaling could play an important role in the segregation of HCV-positive cells into localized clusters, preventing further spread by protecting cells in a neighborhood of the cluster [44]. In another work, *in vitro* experiments have shown that IFN-suppressing wild-type vesicular stomatitis virus (VSV) is out-competed by mutants lacking IFN-suppression when either host-cell IFN response or monolayer spatial structure is removed. However, when monolayer spatial structure is preserved, the IFN-suppressing phenotype emerges as the dominant strain universally [45], emphasizing the importance of spatial structure in determining the effectiveness of the IFN signaling.

The IFN response to virus spread among cells at the host level has clear parallels in infectious disease transmission among individuals at the epidemiological level. First, autocrine signaling has an epidemiological parallel to testing and self-isolation in epidemiological control, where infectious individuals self-isolate in response to becoming aware of their own infection status through testing. In both cases, an individual cell or person’s infectivity is modulated in response to the discovery of their own infection status. Second, paracrine signaling is in a clear analogy to contact tracing and quarantine, where the aim is to trace the at-risk individuals who contacted the infected individual and reduce the risk of further transmission [46]. Third, when the viral spread is mostly spatial, we showed that the collective host response through IFN diffusion leads to an outer layer of protected cells to isolate the infected cells from other susceptible cells. This is a pattern reminiscent of ring vaccination or ring culling [31, 32, 30]. Fourth, IFNs act as communication molecules to signal to neighboring cells is also similar to the spread of disease awareness at the epidemiological scale. As analyzed in Ref. [47], information about a disease spreads to those close to infected individuals in a contact network, and thus decreases the susceptibility of the informed to infection, suppressing the spread of the thee disease.

Overall, given that the IFN response is a highly effective immediate response employed by host cells in a wide variety of tissues and body compartments [37, 8], we reason that it is likely to be a highly effective and robust strategy to prevent virus spread in a host, irrespective of the molecular details of the infection. At the epidemiological level, interventions discussed above, i.e. testing, isolation, contact tracing, ring vaccination/culling as well as spread of awareness, are likely to be effective and robust strategies against the spread of infectious diseases, although their relative effectiveness may depend on how the pathogen spreads through a population. Altogether, further experimental and modeling works on a quantitative understanding of the IFN response against virus infection will continue to offer new insights into virus infection, treatment and control at both the within-host level and the population level.

## Data Availability

All data is available in the manuscript.

## Author contributions

GML, BS and RK conceived the project. GML, HR and RK developed models. GML and HR performed analysis. GML, BS and RK wrote the paper.

## Acknowledgment

We thank Katia Koelle, Rob de Boer, Christopher Brooke and members of the Ke lab for helpful discussions.

## Supplementary Materials

### 5.1 Derivation of *R*_0_ for ODE System

We will derive an expression for the *basic reproductive number R*_0_ for the ODE system (1) via construction of the Next Generation Matrix (NGM) as outlined in ([36]). Since the dynamics of protein and virion production are faster than those governing the cell variables, we will consider the non-cell variables *V* and *F* to be immediately equilibrate (cite Perelson). This yields the following expressions for the equilirbium values of *V* and *F*:

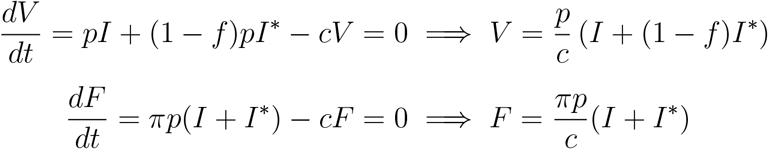

By substituting these equilibrium values into the original system, we are left with the *quasi-equilibrium system* consisting only of cell species shown below.

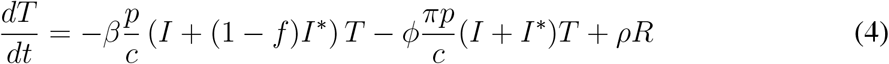

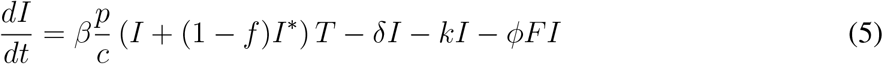

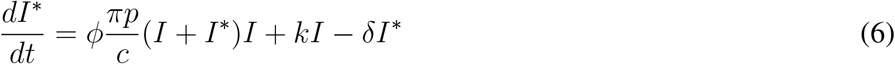

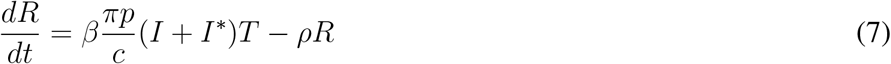

It is important that the NGM analysis be performed on the system in which all state variables are cell species such that the interpretation of *R*_0_ as the number of secondary infected cells caused by the first infected cell remains valid. We now consider the “infectious subsystem” consisting of consisting of equations (5) and (6) above. We compute the Jacobian of this subsystem, evaluating it at the disease-free equilibrium [*T, I, I*^∗^, *R, V, F*]^*T*^ = [*T*_0_, 0, 0, 0, 0, 0]^*T*^, which yields the following:

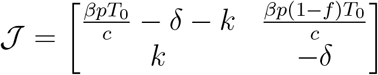

The Jacobian matrix 𝒥 is then partitioned as 𝒥 = **T** + Σ, where the transmission matrix **T** represents epidemiological birth events such as new infections while Σ represents exponentially distributed transition events such as cell death and autocrine-mediated transition from *I* to *I*^∗^.

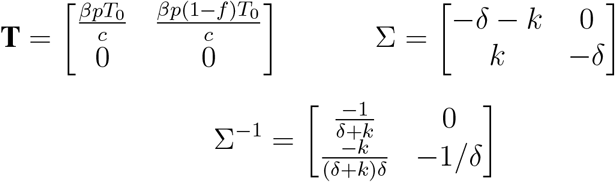

The Next Generation Matrix **K** is then defined to be **K** = − **T**Σ^−1^, and *R*_0_ is defined to be the spectral radius of the **K**. We note here that Σ is always non-singular, as the death of infected cells *I* and *I*^∗^ is required for *R*_0_ to be finite. The following computations were performed with Maple.

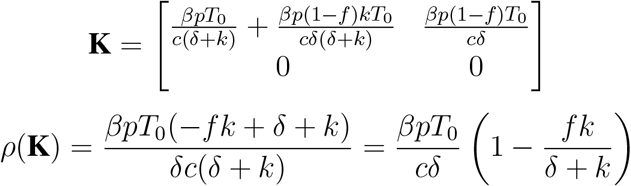

We note that the value of *R*_0_ obtained from the NGM analysis of the quasi-equilibrium system is consistent with the probabilistic interpretation of the terms of the original model, as outlined below:

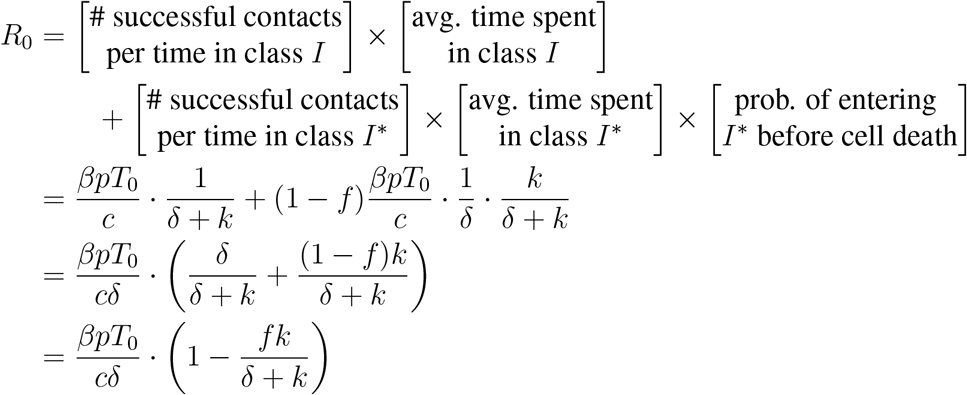

### 5.2 Approximating Peak Viremia in ODE model

We will approximate the peak viral load in consideration of an ODE model that considers only the protective effect of IFN signaling on susceptible cells and not the dampening of virion output from infected cells (i.e., no I* class). Since paracrine IFN signaling is shown to be less influential than autocrine signaling in the ODE context, we consider this model as a “best-case scenario” for the efficacy of paracrine IFN signaling in a spatially-homogeneous setting. This reduces the model (1) to the following system of ODEs:

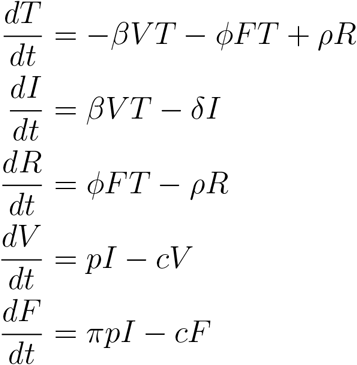

We will once again assume that the compartments *V* and *F* are near equilibrium, so we will take the following approximation in our analysis:

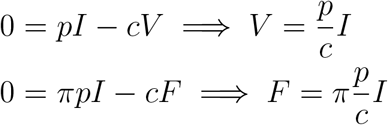

We now consider an approximate model for the cell compartments alone that employs the rapid equilibration of the *V* and *F* compartments. We furthermore assume that in the initial stage of infection the target cell population remains close to the initial value *T*_0_ and that the transition from the refractory class back to the target population is negligible during initial infection (i.e., *ρ* = 0). Furthermore, we take the contact rates for borth both virions and IFNs to be *β*. Since there is mutual unidentifiablity between *p* and *beta* and between *πp* and *ϕ*, we can let *β* = *ϕ* as the parameter *π* can implicitly account for the difference in these contact rates as might be present in biology. Together these assumptions yield the following pair of ODEs:

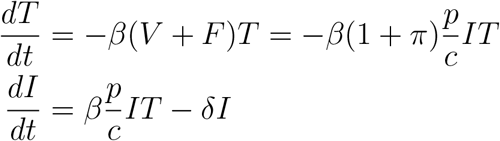

We will now approximate the peak infected cell population. At peak infection, 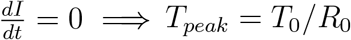 where 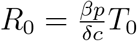. We consider now a phase plane solution to the following:

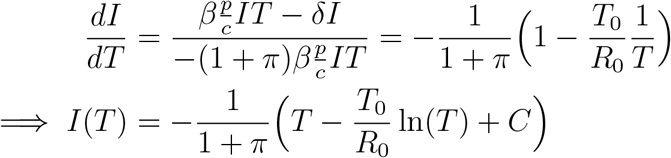

Under the assumption that there is no initial infected cell population,

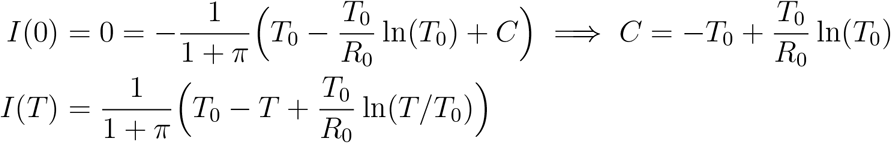

Since we now have *I* expressed explicitly as a function of *T*, we can substitute in the value of *T*_*peak*_ calculated previously:

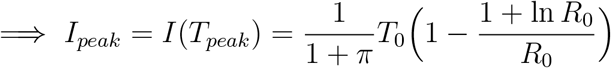

We lastly return to our assumption that under rapid equilibration 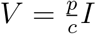, so our expression for the peak viral load is

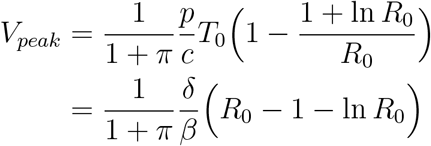

Since *π* = 0 corresponds to the case of no paracrine **IFN** signaling, we see that paracrine **IFN** signaling has the effect of reducing peak viremia by a factor of 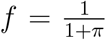 compared to the case of no paracrine **INF**.

### 5.3 Approximating Time to Peak

We will find an approximate expression for the time to peak viremia. We again make the quasi-steady state assumption in *V* and *F*, take *f* = 1 so that autocrine signaling completely shuts down virion output.

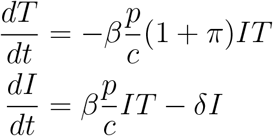

We see that peak infection (*dI*/*dt* = 0) occurs at such a time that *T* (*t*_*peak*_) = *T*_0_/*R*_0_, where 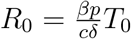. Approximating growth to be exponential until that time, we have:

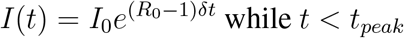

Substituting this expression into the equation for *T*, we can easily solve the single ODE.

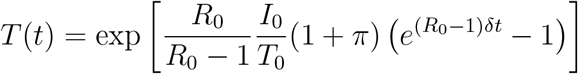

taking *T* (*t*_*peak*_) = *T*_0_/*R*_0_ in the above approximate solution for *T* (*t*), we can easily solve for *t*_*peak*_ and find the following approximate expression.

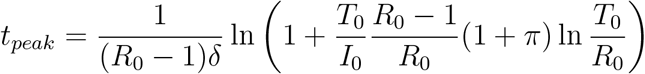

### 5.4 Choice of parameter values

In our models, parameter values were taken or derived mostly from studies of influenza infection. In the ODE model (see Table S1 for parameter values), we set the initial target cell population, *T*_0_ to 4.0 ×10^8^ cells according to a previous estimate of number of target cells for influenza infection [23]. The death rate of infected cells is set to 4/day according to Ref. [23]. The rate of infected cells to become cells in an antiviral state, *k*, is set to 2/day, because Rand et al. [37] showed that cells turn on interferon signaling between 12 to 24 hours post infection. Virion production is set to a rate used in a previous influenza study [33]. Virion clearance rate and the rate that refractory cells become target cells again are set to values estimated by Pawelek et al. [25]. We set the value of *β* such that the viral load peaks around 2 days post infection as seen in the data from Ref. [23]. The value of ϕ is unknown in general. Here we set it to be the same value as *β*, such that the value of *π* represent the relative impact of IFN signaling on cells as compared to the viral infection process.

In the PDE model (see Table S2), we model infection from the upper and the lower respiratory track as a one-dimensional spreading process, and the length of the one-dimensional space is assumed to be 30cm [48]. Therefore, the target cell density is calculated as 4.0 ×10^8^/30 = 1.3 × 10^7^ cells/cm^2^. We also assumed the diffusion coefficient for IFN is much higher than viruses according to Ref. [33] and set *D*_*F*_ = 40 × *D*_*V*_. The value of *β* is increased to ensure that virus population grows over time. Other parameters are kept the same to be consistent with the ODE model.

Overall, the choice of parameter values in our model are consistent with estimates of acute infections in the literature. The overall conclusion of our study is robust to variations of these parameter values within biological plausible range.

### 5.5 Traveling Wave Analysis and Derivation of Wave Speed

We will perform a traveling wave analysis of the reduced PDE model without IFN response given by the following equations:

**Table S1:** Description of parameters in the ODE model.

| Parameter | Description | Units | Default Value | Source |
| --- | --- | --- | --- | --- |
| $T_0$ | initial target cell population | cells | $4.0 \times 10^8$ | [23] |
| $\beta$ | virus contact rate | cm particles<sup>-1</sup> day<sup>-1</sup> | $5 \times 10^{-10}$ | |
| $\phi$ | IFN contact rate | cm particles<sup>-1</sup> day<sup>-1</sup> | $5 \times 10^{-10}$ | |
| $\rho$ | reversion from $R$ to $T$ | day<sup>-1</sup> | 1 | [25] |
| $\delta$ | infected cell death rate | day<sup>-1</sup> | 4 | [23] |
| $k$ | autocrine transition rate | day<sup>-1</sup> | 2 | [37] |
| $f$ | autocrine efficacy | unitless | 0.9 | |
| $p$ | virion production | virion cell<sup>-1</sup> day<sup>-1</sup> | 2400 | [33] |
| $\pi$ | IFN production rate relative to virus production | unitless | 1 | |
| $c$ | virion clearance | day<sup>-1</sup> | 14 | [25] |

**Table S2:** Description of parameters of the PDE model.

| Parameter | Description | Units | Default Value | Source |
| --- | --- | --- | --- | --- |
| $T_0$ | initial target cell density | cells cm<sup>-1</sup> | $1.3 \times 10^7$ | [23, 48] |
| $\beta$ | virus contact rate | cm particles<sup>-1</sup> day<sup>-1</sup> | $7.3 \times 10^{-9}$ | |
| $\phi$ | IFN contact rate | cm particles<sup>-1</sup> day<sup>-1</sup> | $7.3 \times 10^{-9}$ | |
| $\delta$ | infected cell death rate | day<sup>-1</sup> | 4 | [23] |
| $k$ | autocrine transition rate | day<sup>-1</sup> | 2 | [37] |
| $f$ | autocrine efficacy | unitless | 0.9 | |
| $p$ | virion production | virion cell<sup>-1</sup> day<sup>-1</sup> | 2400 | [33] |
| $\pi$ | IFN production rate relative to virus production | unitless | 1 | |
| $c$ | virion clearance | day<sup>-1</sup> | 14 | [25] |
| $D_V$ | virion diffusion | cm<sup>2</sup> day<sup>-1</sup> | 1 | |
| $D_F$ | IFN diffusion | cm<sup>2</sup> day<sup>-1</sup> | 40 | |

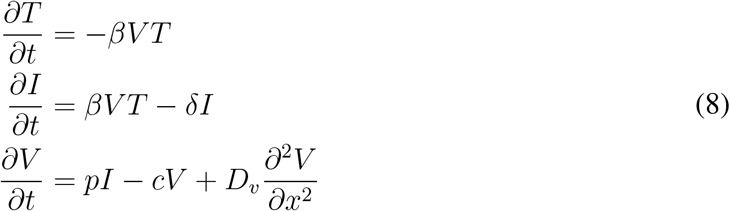

We will consider a traveling wave solution of the system. For each compartment *u*_*i*_(*x, t*) there exists a function *U*_*i*_(*z*) where the solution can be expressed as *u*_*i*_(*x, t*) = *U* (*x* + *vt*). Now for each compartment *u*_*i*_ we make the following substitutions:

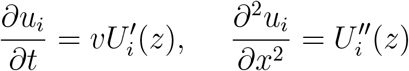

Making the above substitutions and re-writing the second-order equation in *V* as two first order equations, we arrive at the following 4 dimensional system system of first order equations:

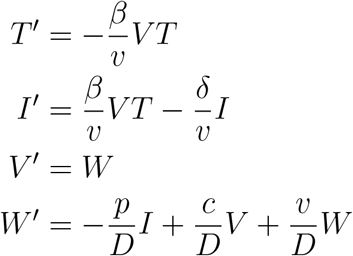

This system has steady state *u*_*e*_ = [*T*_0_, 0, 0, 0]. Evaluating the Jacobian of the system at *u*_*e*_

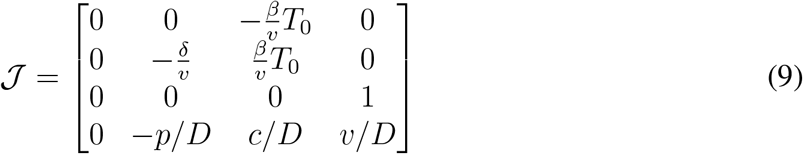

To exclude biologically irrelevant solutions to the system, we will enforce that the eigenvalues of the Jacobian be non-complex to avoid oscillation of the populations about zero. That is to say, the minimum admissible wave speed will occur for the critical value *v*^∗^ at which the the characteristic polynomial of the Jacobian takes a double real root. This of course is the threshold value before which the polynomial takes complex conjugate-pair roots. The block diagonal structure of the Jacobian reveals there to be a zero eigenvalue, so we can examine instead the characteristic polynomial of the full-rank 3 *×* 3 block.

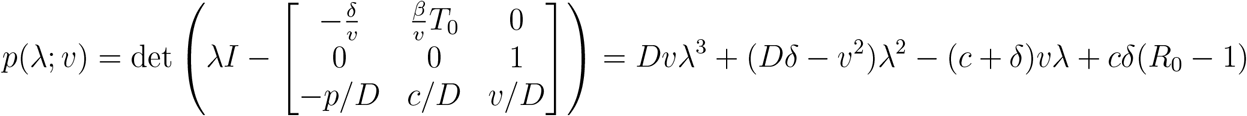

We note the following when *R*_0_ > 1:

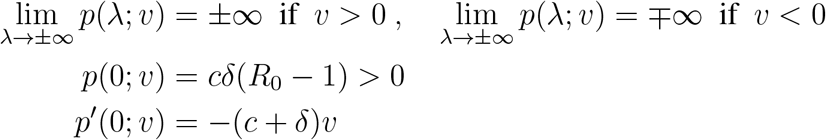

These three observations together ensure that *p*(*λ*) always has one negative real root and attains an extremum for some *λ* > 0. We will now construct *v*^∗^ such that there exists a positive real double root on *R*^+^. For a positive double root to exist it must also be an extremum of the polynomial. This means we solve the following system of equations for *λ* and *v*:

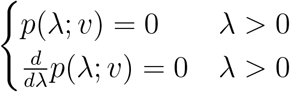

For a given parameter set, this can be done numerically to find a prediction of the traveling wave speed.

### 5.6 Discriminant Method for Determining Exact Expression for Wave Speed

With any polynomial *p*(*λ*) = *Wλ*^3^ +*Xλ*^2^ +*Y λ* +*Z* of degree 3 we can associate a discriminant ∆_3_ = *X*^2^*Y* ^2^− 4*WY* ^3^− 4*X*^3^*Z* −27*W* ^2^*Z*^2^ + 18*WXY Z*.

The minimum admissible traveling wave speed of the system (8) is found to be the critical value *v*^∗^ for which the characteristic polynomial of the Jacobian takes a double root, which is precisely where ∆_3_ = 0. In terms of the discriminant, we frame the problem as:

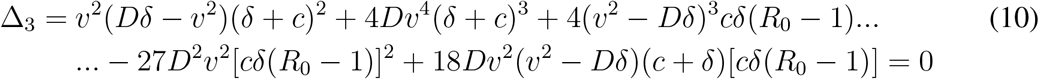

For the simpler problem where there is no death of infected cells and thus the total number of cells is conserved we can further reduce the dimensionality of the problem:

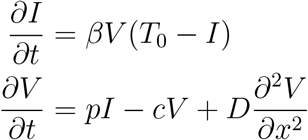

We find the corresponding 1^*st*^ order traveling-wave ODE to have jacobian about the zero equilibrium.

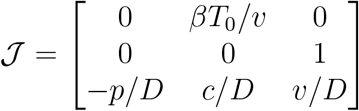

We will consider the discriminant of the characteristic polynomial of the above jacobian matrix to be a function of the wavespeed *v* and collect terms accordingly:

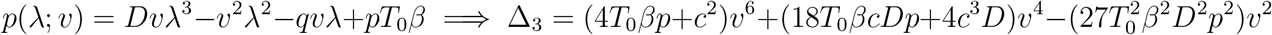

We see that the discriminant here is a sixth order polynomial in *v* with no odd order terms and no constant term. Since the trivial wavespeed *v* = 0 is not of interest, we can neglect the double root of *v* = 0 and look at the following:

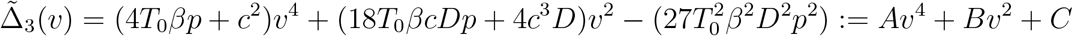

The minimal wavespeed *v*^∗^ will be the largest value of *v* for which 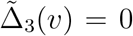. Since the above is quadratic in *v*^2^, it is easily solved by the quadratic formula. This furthermore implies that each positive wave speed admitted also admits an equal and opposite wave speed, as the system is non-advective and thus has no directional bias. Letting *R* = *T*_0_*βp*, we have the following expression for the wave speed:

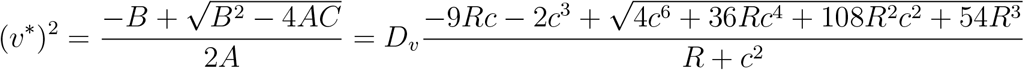

Recalling the parameter values used from Table S2, we can choose smaller order terms to neglect to arrive at a simpler expression that is sufficiently accurate for a neighborhood of our parameter set. Here, observing *R* ≈ 10^4^ and *c* ≈ 2 *×* 10^1^, we arrive at the following reduced expression:

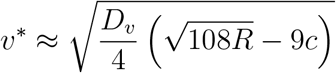

This value is shown to be consistent with traveling speeds measured from numerical simulations (*relative error* ≈ 5%). Since this method appears effective in producing an accurate closed form expression for the wave speed, we wish to recycle this logic as much as possible. For the more general case where *δ* ≠ 0, we would like to make small simplifications that will eliminate odd order terms and allow use of the quadratic formula. Specifically we will make the assumption that *v*^2^ >> *Dδ* and will substitute *X* = (*Dδ* →*v*^2^) → −*v*^2^. Under this substitution, the non-zero positive root of the discriminant is approximated by quadratic formula and found be consistent with numerical simulation (*relative error* ≈ 2 − 5%). The closed form is very complicated and is not included here.

### 5.7 Cellular Automata Model

An infected cell produces `virus_prod` virions per time step. These are the “successful” virions, which are the virions that successfully contact another cell before they can be degraded or otherwise cleared. In this sense, the production number `virus_prod` is considerably smaller than the actual quantity of virions that would be produced by the cell in a single time step. The nominal values of the CA parameter values may be found in Table S3

Rather than allowing the virions produced to diffuse along the lattice explicitly, for each virion produced we select a recipient cell according to the following stochastic process. We determine a time-to-contact, ∆*t*, where ∆*t* is drawn from an exponential distribution with parameter *λ*, chosen such that 1/*λ* is the average time for a virion to reach a recipient cell. Then a distance *r* is drawn from a normal distribution with mean 0 and standard deviation 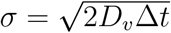, where *D*_*v*_ = `virus_diff` is the virion diffusion coefficient. An angle *θ* is then drawn from a uniform distribution *𝒰* (0, 2*π*), and the cell found at a distance *r* in the *θ* direction from the producing cell is chosen to receive the virion. That is, for a producing cell located at position (*x*_0_, *y*_0_), a recipient cell location (*x, y*) is chosen as follows:

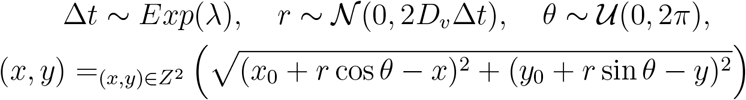

This procedure is equivalent to sampling the distribution given by the solution of the Diffusion Equation *V*_*t*_ = *D*_*v*_∆*V* with Dirac-delta initial condition evaluated at time *t* = ∆*t*. After being selected as the recipient of a virion, a cell enters the exposed state if it is not already exposed, productively infected, or protected. After `virus_prod_delay` time steps, the exposed cell becomes productively infected and begins to produce virions and will continue to do so for the remainder of the simulation. Similarly, after `ifn_prod_delay` time steps from exposure, the exposed cell begins to produce IFN particles, and continues to for the remainder of the simulation. We assume the delay in IFN production to be smaller than the delay in virion production. IFN particles are produced and transfected instantaneously into recipient cells in the same way as virions, with separate production and diffusion parameters, `ifn_prod` and `ifn_diff`, respectively.

**Table S3:** Parameters of the CA model.

| Parameter | Description | Units | Default Value |
| --- | --- | --- | --- |
| `virus_prod` | virus production rate | virus step<sup>-1</sup> | 1 |
| `ifn_prod` | IFN production rate | IFN step<sup>-1</sup> | NA |
| `virus_diff` | virus diffusion coeff. | cell-width<sup>2</sup> step<sup>-1</sup> | 0.2 |
| `ifn_diff` | IFN diffusion coeff. | cell-width<sup>2</sup> step<sup>-1</sup> | 1 |
| `virus_prod_delay` | post-exposure wait time | steps | 14 |
| `ifn_prod_delay` | post-exposure wait time | steps | 8 |

The parameters of the CA model were selected to reflect experimentally verified values, though it must be noted that experimental conditions may not adequately represent the in-host environment. `virus_prod` is taken to have unit value, implying that time unit `step` is defined to be the average time for an infected cell to produce a single viral progeny. Taking the virion production parameter from the DE models *p* = 2400, then we can interpret `step` = 1/2400day. Since the spatial unit in the CA model is one cell width, by taking the diffusivity of an influenza virion in human periciliary fluid to be 10^−12^ [48] and the average diameter of an epithelial cell to be 12.7*µ*m [49], we can approximate the virion diffusivity in our units to be approximately 0.2. The IFN diffusivity `ifn_diff` has not been experimentally studied, so is taken here to be up to an order of magnitude higher than that of the virion due to the difference in particle size. This is likely an influential parameter that could be a key object of future study.

## Supplementary Figures

**Figure S1:**
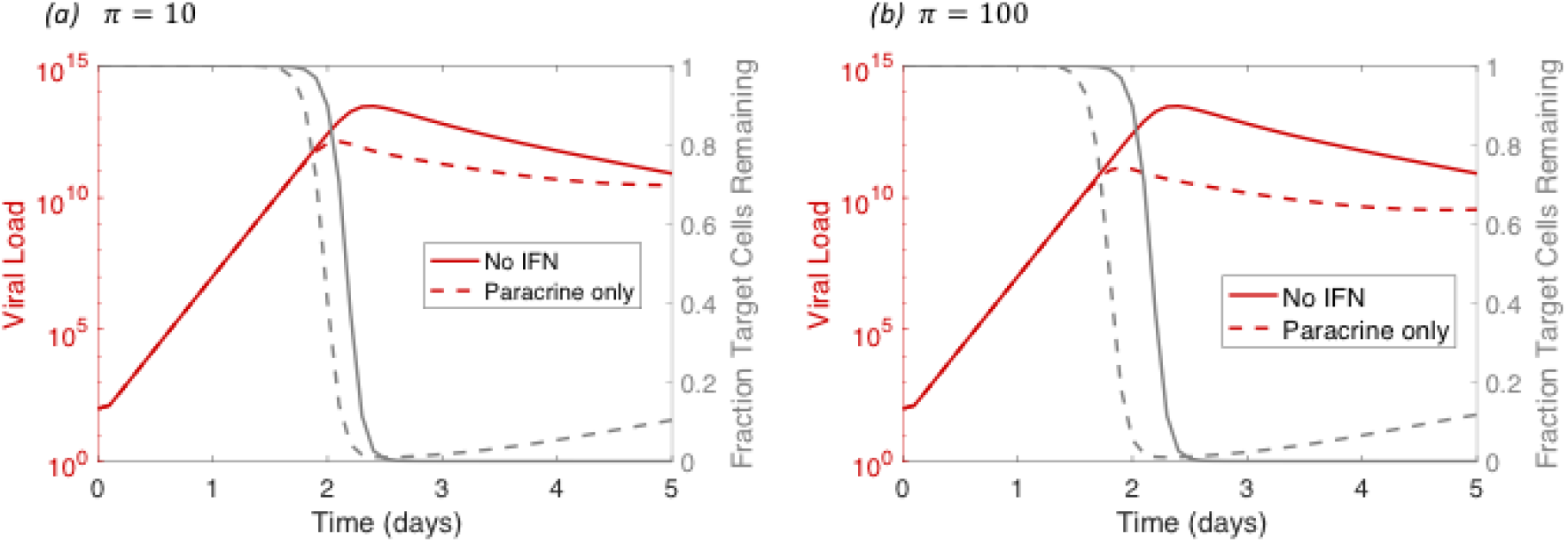
Even supraphysiologial levels of IFN production *π* do not affect initial exponential growth rate. Very high levels of free IFN can shorten the time to peak viremia, but do not demonstrate the ability to halt establishment of systemic infection.

**Figure S2:**
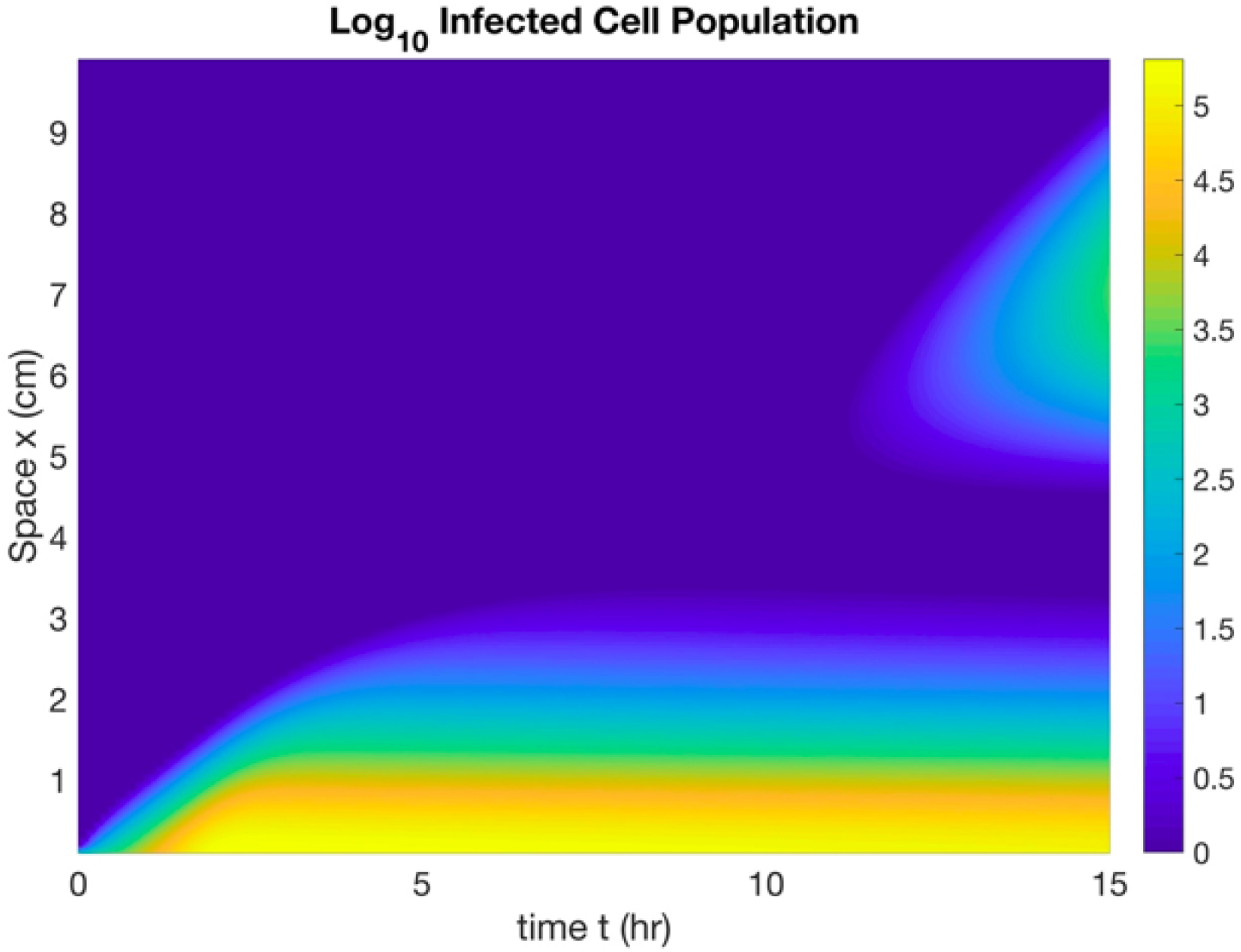
Given sufficient time, infection will reemerge despite strong IFN signaling. Shown is a representation of log_10_ *I*(*x, t*) over sufficient time scale for infection to reemerge far from initial location. This phenomenon is an artifact of the continuum approximation inherent in the PDE fomulation of the model. Due to the parabolic nature of the PDE, compact data becomes immediately non-zero in the entire domain, leading to inevitable reemergence of infection in locations where only fractional amount of virus is present.

